# Major new lineages of SARS-CoV-2 emerge and spread in South Africa during lockdown

**DOI:** 10.1101/2020.10.28.20221143

**Authors:** Houriiyah Tegally, Eduan Wilkinson, Richard R Lessells, Jennifer Giandhari, Sureshnee Pillay, Nokukhanya Msomi, Koleka Mlisana, Jinal Bhiman, Mushal Allam, Arshad Ismail, Susan Engelbrecht, Gert Van Zyl, Wolfgang Preiser, Carolyn Williamson, Francesco Pettruccione, Alex Sigal, Inbal Gazy, Diana Hardie, Marvin Hsiao, Darren Martin, Denis York, Dominique Goedhals, Emmanuel James San, Marta Giovanetti, Jose Lourenco, Luiz Carlos Junior Alcantara, Tulio de Oliveira

## Abstract

In March 2020, the first cases of COVID-19 were reported in South Africa. The epidemic spread very fast despite an early and extreme lockdown and infected over 600,000 people, by far the highest number of infections in an African country. To rapidly understand the spread of SARS-CoV-2 in South Africa, we formed the Network for Genomics Surveillance in South Africa (NGS-SA). Here, we analyze 1,365 high quality whole genomes and identify 16 new lineages of SARS-CoV-2. Most of these unique lineages have mutations that are found hardly anywhere else in the world. We also show that three lineages spread widely in South Africa and contributed to ∼42% of all of the infections in the country. This included the first identified C lineage of SARS-CoV-2, C.1, which has 16 mutations as compared with the original Wuhan sequence. C.1 was the most geographically widespread lineage in South Africa, causing infections in multiple provinces and in all of the eleven districts in KwaZulu-Natal (KZN), the most sampled province. Interestingly, the first South-African specific lineage, B.1.106, which was identified in April 2020, became extinct after nosocomial outbreaks were controlled. Our findings show that genomic surveillance can be implemented on a large scale in Africa to identify and control the spread of SARS-CoV-2.

## Main text

Severe Acute Respiratory Syndrome Coronavirus 2 (SARS-CoV-2) is a novel betacoronavirus, first detected in China in December 2019^1,2^. Since then, the coronavirus disease (COVID-19) has developed into a global pandemic, resulting in several waves of epidemics around the world, infecting nearly 30 million people, and causing > 900 thousand death**s** by 15 September 2020^3^. Lockdown and travel restriction measures have varied from country to country, dictating the profile of local epidemic outbreaks. Through the unprecedented sharing of SARS-CoV-2 sequences during this pandemic, including from one of the first cases in Wuhan, China (*MN908947.3*)^2^, genomic epidemiology investigations globally are playing a major role in characterizing and understanding this emerging virus^4–9^. SARS-CoV-2 has typically been classified into two main phylogenetic lineages, lineage A and lineage B. While both lineages originated in China, lineage A spread from Asia to the rest of the world, whereas lineage B predominantly spread from Europe, both circulating widely around the world^10^.

## Near-real-time genomic monitoring of SARS-CoV-2 transmission

The COVID-19 epidemic in South Africa is by far the biggest in Africa, with > 600,000 individuals infected and > 15,000 deaths by mid-September 2020. The first case of SARS-CoV-2 infection in South Africa (SA) was recorded in KwaZulu-Natal (KZN) on 5 March 2020 in a returning traveler from Italy. Around mid-March, cases of community transmission were reported across the country. The profile of SARS-CoV-2 epidemiological progression in South Africa was largely influenced by the implementation of lockdown measures in the early phases of the epidemic and the subsequent easing of these measures. On 26 March, the government-imposed nation-wide lockdown included the prohibition of all gatherings, travel restrictions, and closure of non-essential businesses and schools^11^. Although the epidemic was growing, lockdown measures were progressively eased on 1 May and on 1 June to mitigate negative impacts on the country’s economy. Restrictions were further relaxed first on 17 August, once the peak of new daily infections had passed, and again on 1 October (Fig 1A). The epidemic in South Africa can generally be characterized by two important phases, one dominated by travel-related “early introductions”, and the second being the period of “peak infections” (Fig 1A).

**Figure 1:**
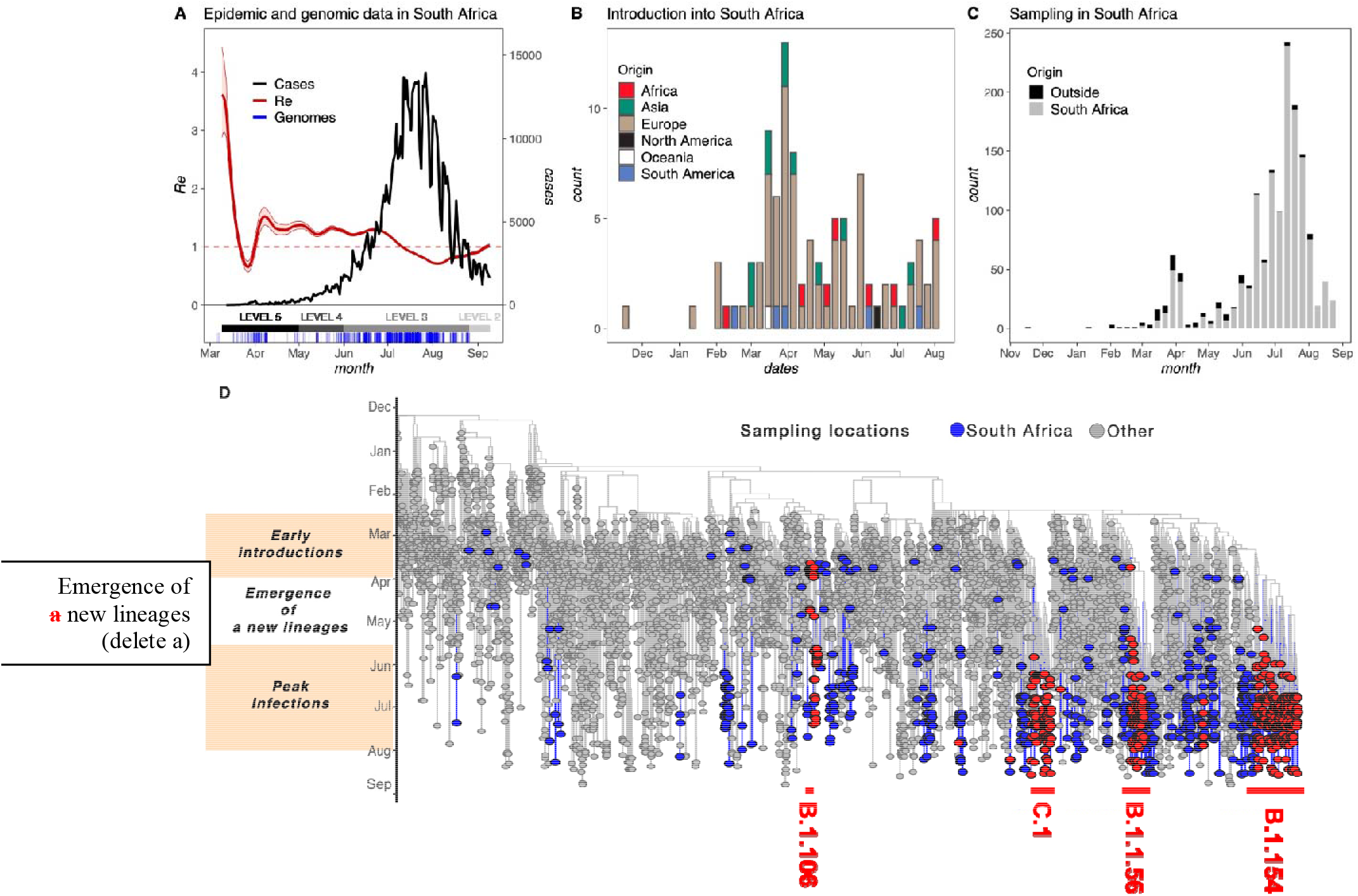
Monitoring SARS-CoV-2 Epidemic in South Africa using genomic sequencing. A) Epidemiological curve showing the progression of daily COVID-19 numbers in South Africa (black), changes in R estimations (red), lockdown levelsand the timing of genomic sampling in South Africa (blue) from the beginning of the epidemic to 15 September. B) Estimated numbers of introductions into SA coloured by region of origin. C) Overall sampling of genomes in South Africa coloured by whether the genomes are associated with introduction events (black) or not (grey). D) Maximum clade credibility (MCC) tree of 7213 global genomes (grey) including 1365 South African sequences (blue), indicating a period of “early introductions” and a period of “peak infections” separated by a period of “emergence of new lineages”. The three largest monophyletic lineage-associated transmission clusters in South Africa are labelled.

We monitored the likelihood of SARS-CoV-2 transmission by estimating the effective reproduction number, R, which provides a measure of the average number of secondary infections caused by an infected person^12^. Typically, a growing epidemic is characterized by R > 1 and R < 1 indicates a slowed progression. At the start of the epidemic, we estimated the R value to be > 3, quickly falling after the start of lockdown to a value of < 1. A subsequent jump in the R value to > 1 was found to be concurrent with the timing of a number of localized outbreaks in the country, including nosocomial outbreaks^13^. The R value again dropped to < 1 at the beginning of August, coinciding with a decrease in the daily number of positive cases recorded (Fig 1A).

Genomic epidemiology is important to understand SARS-CoV-2 evolution and track the dynamics of transmission across the world^4–9^. By 15 September 2020, at the tail end of the epidemiological peak in the country, we had produced 1365 high-quality SARS-CoV-2 whole genomes (>90% coverage; publicly shared on GISAID^14^) in our laboratories as part of the Network for Genomic Surveillance (NGS-SA) consortium^15^. These whole genomes were sampled in eight of the nine provinces of South Africa and in all the districts of KwaZulu-Natal province, (Supplementary Fig S1), and represented consistent sampling from the beginning of the epidemic and corresponding to important events of the epidemiological progression (Fig 1A).

We estimated maximum likelihood (ML) and molecular clock phylogenies for a dataset containing 7213 global genomes, including 1365 South African genomes, sampled from 24 December 2019 to 26 August 2020 (Fig 1C). Time-measured phylogeographic analyses estimated at least 101 introductions into South Africa. The bulk of important introductions happened before lockdown from Europe, where the epidemic was most quickly progressing at that time (Figure 1B). Although at least 67 introduction events are inferred to have occurred after lockdown, these represent only 5% of the genomes that were sampled following lockdown (Fig 1C). In the early phases of the epidemic, before April, 34 introductions were inferred from 35 genomes sampled (97.1%), which we call “early introductions” (Fig 1B). The small number of apparent introductions after lockdown can be explained by more intensive genomic sampling at later stages, which likely revealed introduction events linked to previously undetected transmission chains.

The early introductions were mostly isolated cases with a few instances of small onward transmission clusters, in contrast with large transmission clusters during the peak infections phase (Fig 1C). The time period between these two phases was inferred to be characterized by localized transmission events which saw the emergence and spread of new lineages, which were later amplified during the peak of the epidemic. The South African genomes in this study were assigned to 42 different lineages based on the proposed dynamic nomenclature for SARS-CoV-2 lineages^10^. This included 16 South Africa specific lineages, defined as being lineages that are presently predominant in South Africa by cov-lineages.org as of 15 September 2020^16^ (Supplementary Fig S2). One of these has been assigned a novel SARS-CoV-2 main lineage classification, lineage C, the parent of which is lineage B.1.1.1.

Extensive SARS-CoV-2 genomic sampling, which spanned the whole duration of the epidemic and increased during its peak, allowed for such lineage emergence to be observed, similar to the UK’s genomic investigation of SARS-CoV-2^17^. We focused on the three largest monophyletic lineage clusters (C.1, B.1.1.54, B.1.1.56,) that spread in South Africa during lockdown and then grew into large transmission clusters during the peak infections phase of the epidemic (Fig 1C). Dominance of the epidemic by novel SA-specific lineages likely happened due to lockdown-imposed travel restrictions fueling a largely local epidemic. Accordingly, these three main lineages account for 42% of all sampled South African sequences. In addition to the three most widespread lineages in South Africa, our analysis also focused on an early lineage, B.1.106, that emerged during nosocomial outbreaks in KZN province. This lineage was responsible for 16% of the infections in KZN at the start of April, but its prevalence decreased as the outbreaks were controlled.

## Spatiotemporal spread and associated mutations of the three main lineage clusters in South Africa

B.1.1.54, B.1.1.56, and C.1 represent the three largest monophyletic clusters associated with South Africa-specific lineages that emerged and spread in the country following lockdown and into the peak of the epidemic. They contain 320, 104, and 151 genomes, respectively, which represents 42.1% of the total genomes in this study (Supplementary Table 1), with a clear over-representation in later stages of the epidemic (Fig 2D). Genomes belonging to these lineages were sampled in five adjacent provinces of South Africa and in all 11 districts of KZN province (Fig 2B, 2C, Supplementary Figure S3), and corresponded to timepoints spanning from April to August 2020 (Figure 2B, 2C). We compared Ct scores for genomes that we generated (n=1122) and show that there is no significant difference between the Ct scores of sequences belonging to these three lineages and the others (Supplementary Fig S4). This suggests that the fast spread of the lineages of interest is likely a result of localized outbreaks and expected transmission dynamics, rather than caused by any fitness advantage.

**Figure 2:**
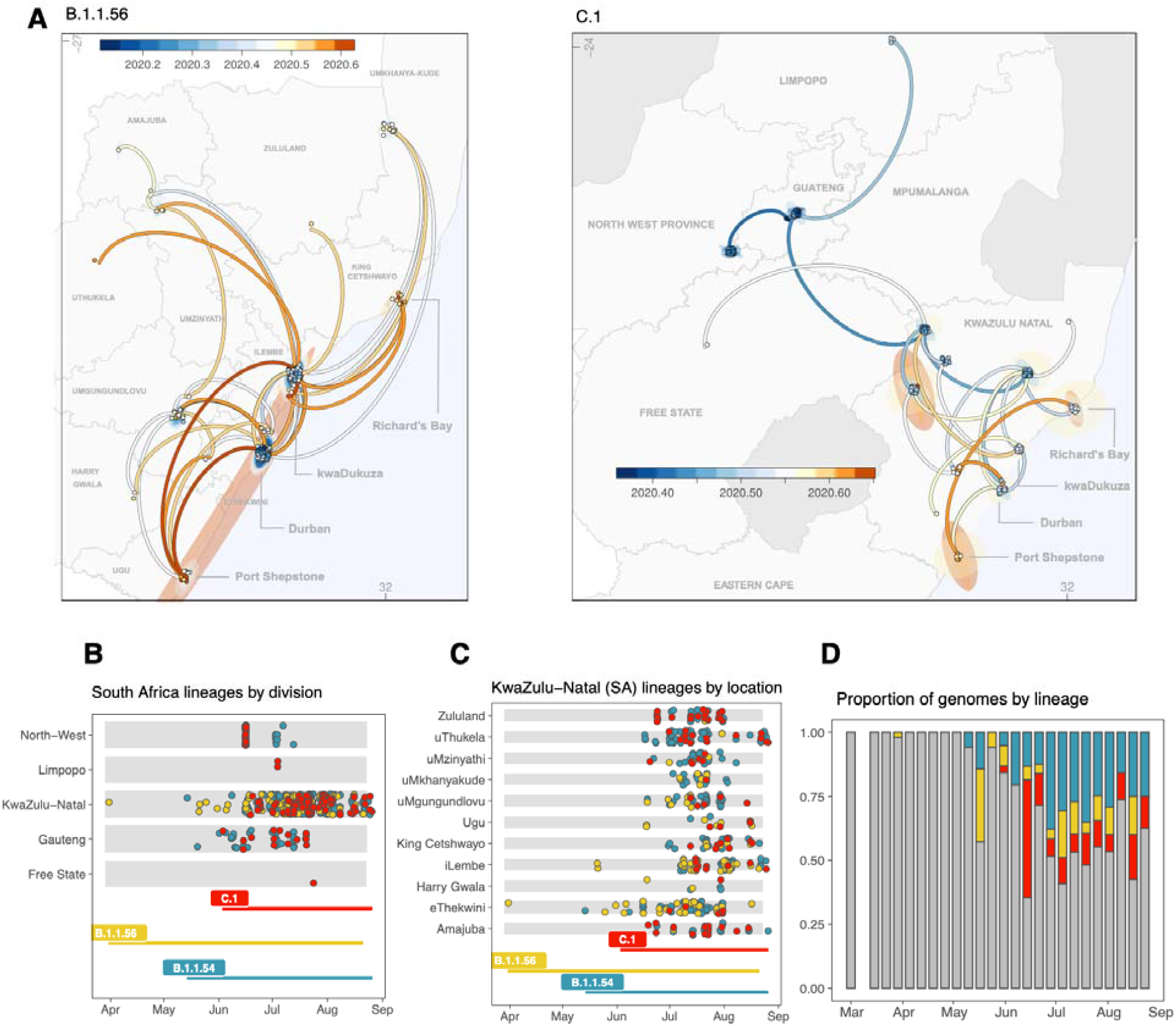
Geographical distribution and spread of lineage clusters in (5) five provinces and all districts of KZN. A) Mapping the spread of the B.1.1.56 cluster (left) and the C.1 cluster (right) from phylogeographic reconstructions. Time scale is specified in decimal dates from 2020.2 (March 2020) to 2020.6 (July 2020). Eastern Cape misspelled – to be fixed in next iteration. B) Sampling timeline and locations of genomes belonging to each lineage cluster in (5) five provinces. C) Sampling timeline and locations of genomes belonging to each lineage cluster in all 11 districts of KZN province. D) The progression of the proportions of genomes belonging to the main lineage clusters over time (B.1.1.54: blue, B.1.1.56: yellow, C.1: red), grey: others).

In order to better understand the spatiotemporal diffusion of South African specific lineages, we used a continuous phylogeographic model that maps the phylogenetic nodes to their inferred geographical origin locations (Fig 2A). Bayesian MCMC analysis in BEAST suggests these lineages emerged during the early phase of the South African epidemic between February and May 2020 (Supplementary Fig S6). Our phylogeographic reconstruction suggests that lineage B.1.1.56 emerged in the city of Durban (eThekwini, ETH) around mid March 2020 (95% HPD Jan – April 2020). It appears that from June onwards, this lineage quickly disseminated throughout KZN to all of the districts. This occurred when the country moved from lockdown level 4 to 3, which allowed greater movement of people and goods between districts. Lineage C.1 most likely emerged in early May 2020 (95% HPD 2020-04-24 – 2020-05-24) in the city of Johannesburg, located in Gauteng province, from where it quickly spread to the adjacent North-West province, where it caused a large nosocomial outbreak^18^. Furthermore, the lineage spread through two independent events to the northern province of Limpopo and to northwestern KZN. From this location, the lineage further spread into all districts of KZN and to the adjacent Free State Province. Unfortunately, lineage B.1.1.54 showed poor temporal signaling (Supplementary Fig S5) and therefore Bayesian spatiotemporal analyses could not be performed for this cluster. A closer look at the cluster (from the ML timetree) is, however, shown in Supplementary Figure S6 and indicates that this lineage was first sampled in KZN and Gauteng and later spread in large numbers in the provinces of KZN, North West and the Free State.

We analysed the sequences of the three main lineage clusters in order to determine respective lineage-defining mutations. On average, sequences in the C.1 cluster accumulated roughly 16 mutations, while B.1.1.56 and B.1.1.54 have approximately 13-14 mutations relative to the Wuhan reference (*NC_045512*) (Fig 3A). This is relatively higher than the number of acquired mutations in other sequences, which is consistent with these three lineages having emerged more recently than the rest, hence accumulating more genomic changes. Sequences are assigned lineages based on the presence of certain lineage-defining mutations (Supplementary Fig S7). The sequences belonging to B.1.1.54, B.1.1.56 and C.1 all have the mutations that define their B.1.1 parental lineage (C.1 was previously known as B.1.1.1.1) (Fig 3B, in black), including the 23493 A>G (Spike D614G) mutation, with additional mutations that differentiate them (Fig 3B, in red). Sequences in B.1.1.54 have the *12503T>C (NSP8: Y138H)* and *29721C>T* mutations in > 90% frequency, similar to *22675C>*T for B.1.1.56, and *4002C>T (NSP3: T428I), 10097G>A (3C-like proteinase: G15S), 13536C>T, 18747C>T* and *23731C>T* for C.1 (Fig 3B). The early hospital-linked lineage B.1.106 was defined by the *16376C>T (helicase: P47L)* mutation. Five of these mutations, *12503T>C, 16376C>T, 18747C>T 29721C>T* and *22675C>T*, are predominantly present in South African genomes, with just a few occurrences in the rest of the world (Fig 3C and Supplementary Fig S8), whereas the rest of the lineage-defining mutations are also common in the rest of the world (Supplementary Fig S8). There are two other high prevalent mutations on the Spike protein in the B.1.56 and C.1. lineages, 22675C>T and 23731C>T, but these are both synonymous mutations.

**Figure 3:**
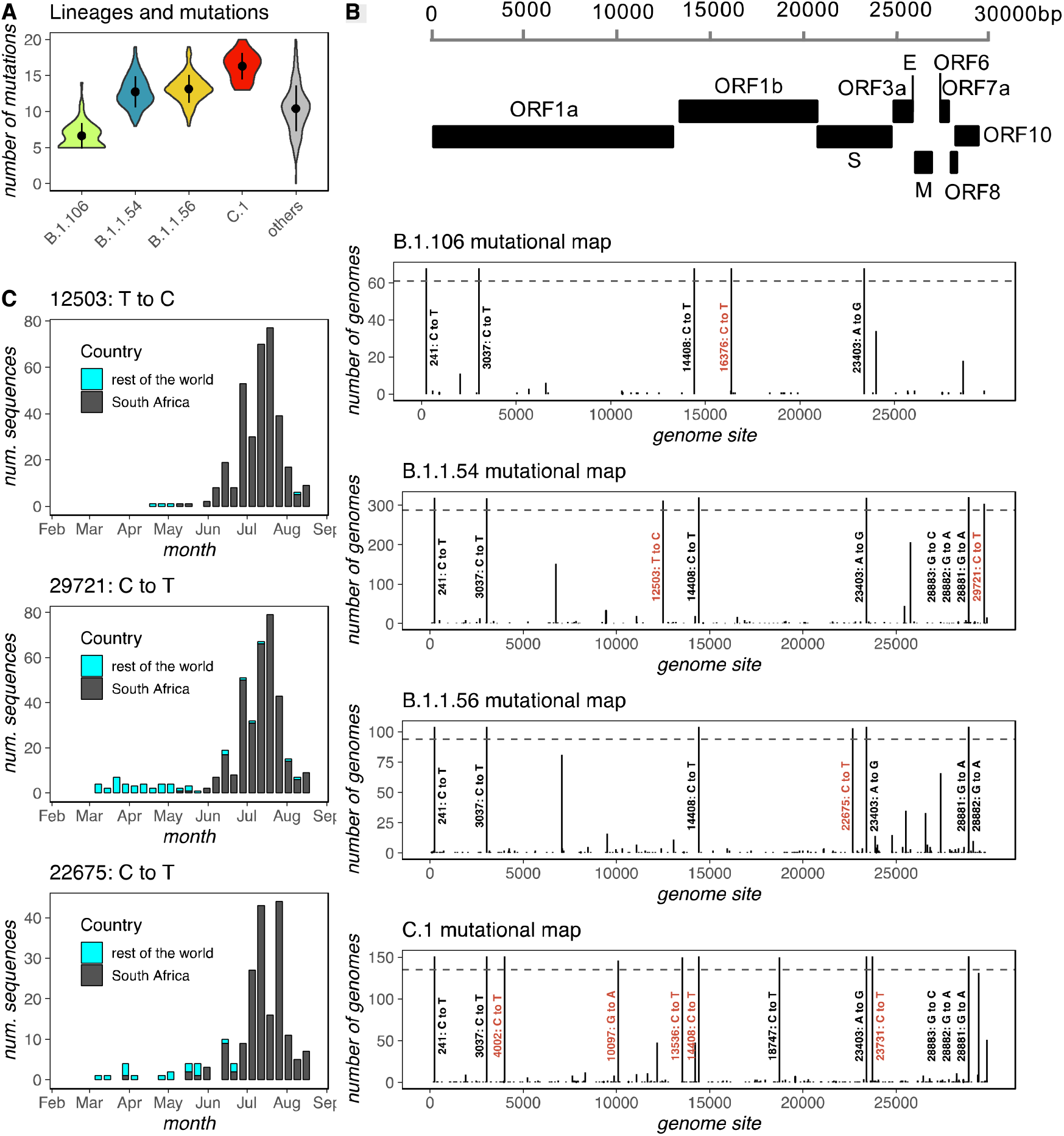
Lineage-defining mutations of the three main SA lineage clusters. A) Violin plot showing the number of mutations in each cluster. B) Variant maps of the most common mutations in each cluster mapped against the SARS-CoV-2 genomes. Most common mutations defined as mutations present in >90% of the genomes in that group (dotted blue line). C) Change in frequency of some unique South African mutations over time in South Africa vs the rest of the world.

Major contributor to linage amplifications in South Africa were hospital outbreaks For example, as previously mentioned, lineage C1 was amplified in a nosocomial outbreak in the North West Province in April 2020^18^ before spreading to KZN and other provinces. Another South African lineage, B.1.106, also emerged in a nosocomial outbreak in KZN in April 2020. This was a large outbreak that infected 100 heath care staff and 65 patients, and dominated most of the early infections in Durban, South Africa (Figure 4B). This nosocomial outbreak attracted national attention as it was responsible for 14% of the infections in KZN and over 45% of the national deaths in early April 2020. We used genetic sequencing, together with active outbreak investigation to understand how the virus entered and spread in this hospital^13^. This lineage also spread to the population and caused a second nosocomial outbreak in a nearby hospital that infected 15 health care workers (Figure 3A). These two nosocomial outbreaks were identified within days of the first infection and were followed with very active infection and prevention control measures^13,19^. The B.1.106 lineage largely subsided following the outbreak investigations and isolation of all infected individuals. The B1.106 lineage’s prevalence at the population level decreased quickly after June 2020 (Figure 4).

**Figure 4:**
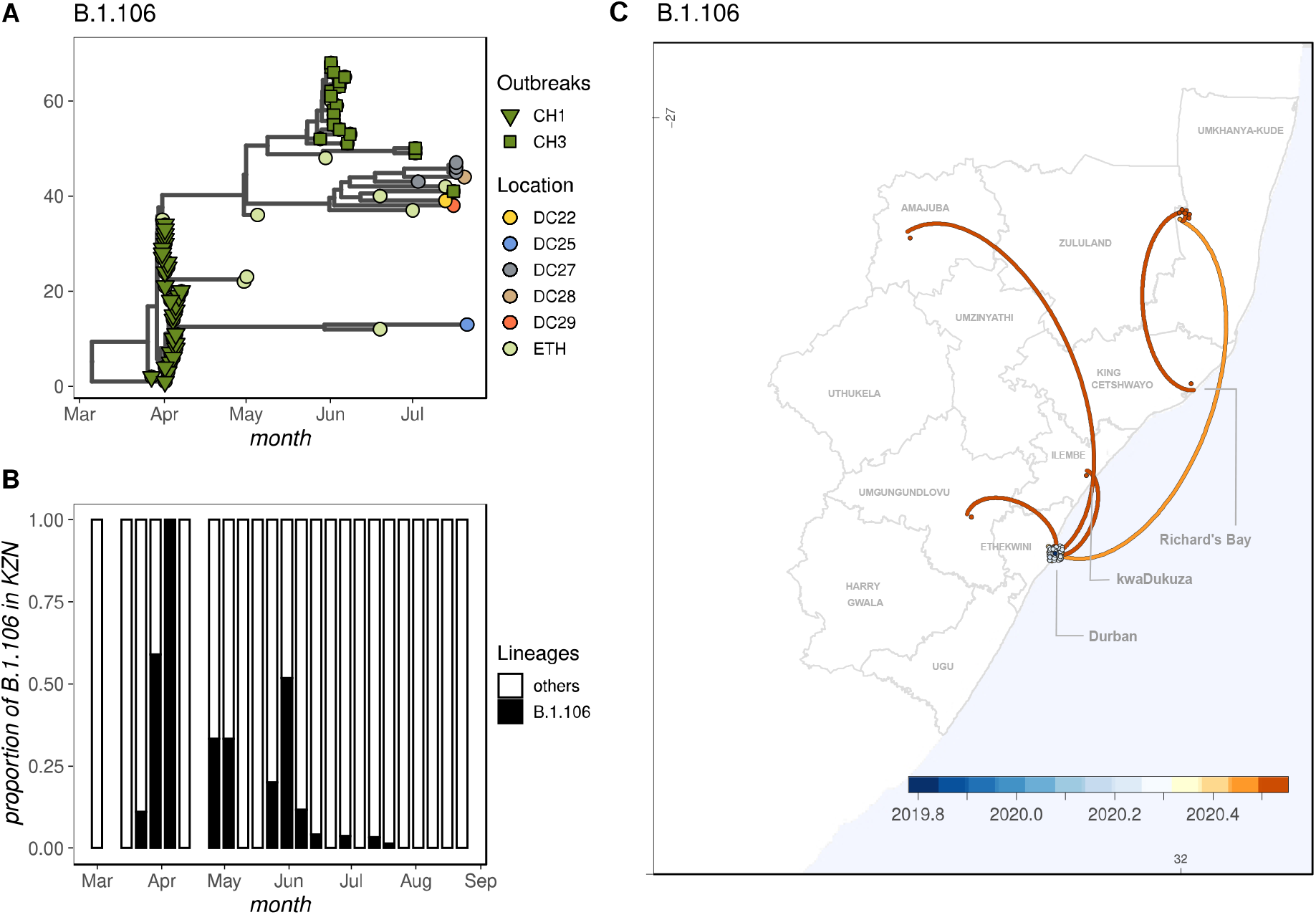
Lineage B.1.106 phylogenetic tree and dispersion throughout KwaZulu-Natal (KZN). A) Phylogenetic tree of B1.1.106 sequences by nosocomial outbreak (Clustered Hospital 1, CH1 and CH3) and district location. B) Proportion of sequences classified as B1.106 over time in KZN. C) Mapping the spread of the B.1.106 lineage from phylogeographic reconstructions. Time scale is specified in decimal dates from 2019.8 (November October 2020) to 2020.5 (June 2020). Shade circular patterns represent confidence intervals of location estimation.

## Discussion

We report an in-depth analysis of the spread of SARS-CoV-2 in South Africa, showing that the bulk of introductions happened before lockdown and travel restrictions were implemented at the end of March 2020. However, despite drastic lockdown measures, the pandemic spread quickly, causing over 600,000 laboratory confirmed infections. In order to track the evolution of the virus, we formed the NGS-SA, a consortium of genomics and bioinformatics scientists that worked with national government laboratories to quickly generate and analyze data in the country. We produced 1,365 SARS-CoV-2 whole genomes and mapped the emergence of 16 South African specific novel lineages. These lineages became established during the hard lockdown and spread widely in the country. We find that three main lineages were responsible for almost half of all the infections in South Africa. Despite a relative sequencing bias in KZN, we were able to detect these major lineages across multiple provinces. It is therefore likely that more extensive sampling throughout the country could pick up the spread of these lineages all over the country, especially as lockdown levels were eased and mobility increased. Indeed, recent data from Cape Town also identified the C1 lineage, which is the most geographically widespread lineage in South Africa.

Genomic data was also used in real-time to identify and control nosocomial outbreaks. For example, the B.1.106 lineage, which was the first South African lineage to be identified, was used to understand how the virus spread inside a large hospital in the country. The lessons learned in this outbreak were used to quickly control a second nosocomial outbreak. The active outbreak investigation itself may have limited the spread of this lineage. Our analysis therefore shows that a number of SARS-CoV-2 lineages, each with unique mutations, emerged within localized epidemics during lockdown even as the introduction of new lineages from outside South Africa was being curbed.

It is currently unknown if any of the mutations originating in South Africa have a fitness advantage in terms of transmission, viral replication, or a reduced immunogenicity in the South African population. That many of the mutations are synonymous and that differences in Ct values do not seem to be appreciably affected by the infecting viral strain argues against selection for fitter variants. It is important to note that the four main lineages in South Africa contain the D614G mutation on the spike gene. We are currently investigating limits to cross-reactivity between strains. Limited cross-reactivity may lead to effects such as antibody dependent enhancement (ADE) in response to a vaccine with a non-native strain. ADE occurs in infections such as Dengue when a previously infected individual is infected with a second strain of virus, which antibodies from the first infection can bind to but not neutralize^20^.

In conclusion, this study further emphasizes the usefulness of integrating genomic surveillance methods to understand SARS-CoV-2 spread in local settings. Furthermore, genomics data can also be used in real-time to inform and consolidate national outbreak investigation and response strategies in Africa.

## Data Availability

All of the sequence data, i.e. 1365 whole genomes of SARS-CoV-2, have been deposited at GISAID.

## Acknowledgements

This research was funded by The South African Medical Research Council (SAMRC), MRC SHIP and the Department of Science and Innovation (DSI) of South Africa. KRISP is funded by a core award of the South African Technology Innovation Agency (TIA). We would like to thank Prof. Andrew Rambaut and Dr. Áine O’Toole for scientific discussion on how to include the South African lineages on PANGOLIN dynamic classification.

