## Supplementary Material for "Major new lineages of SARS-CoV-2 emerge and spread in South Africa during lockdown"

**Supplementary section - Materials and Methods**

**Contents:**

- Materials and Methods

**Additional Supplementary materials:**

- Supplementary Figures
- Data S1 – South Africa dataset metadata (Excel file)
- Data S2 – Acknowledgements to sequence-generating laboratories in South Africa dataset (PDF file)
- Data S3 – Acknowledgements to sequence-generating laboratories in global reference dataset (Excel file)

Ethical statement

We obtained deidentified remnant nasopharyngeal and oropharyngeal swab samples from patients testing positive for SARS-CoV-2 by RT-qPCR from public health and private medical diagnostics laboratories (Data S1). The project was approved by University of KwaZulu-Natal Biomedical Research Ethics Committee. Protocol reference number: BREC/00001195/2020. Project title: COVID-19 transmission and natural history in KwaZulu-Natal, South Africa: Epidemiological Investigation to Guide Prevention and Clinical Care. This project was also approved by University of the Witwatersrand Human Research Ethics Committee. Clearance certificate number: M180832. Project title: Surveillance for outpatient influenza-like illness and asymptomatic virus colonization in South Africa. Sequence data from the Western Cape was approved by the Stellenbosch University HREC Reference No: N20/04/008_COVID-19. Project Title: COVID-19: sequencing the virus from South African patients.

Epidemiological data

We analyzed COVID-19 cases counts in South Africa from publicly released data up to 15th September 2020 from the National Department of Health (NDoH) and the National Institute for Communicable Diseases (NICD) in South Africa. This was accessible through the repository of the Data Science for Social Impact Research Group at the University of Pretoria (<https://github.com/dsfsi/covid19za>)^1^. The NDoH releases daily updates on the number of new confirmed cases, deaths and recoveries, with a breakdown by province. For correlation with government epidemic control measures, we information from government press releases and speech transcripts was extracted. To illustrate the epidemic progression, the daily number of confirmed cases for South Africa was plotted alongside a timeline of lockdown levels and variation in estimated virus reproduction number until the 15th of September 2020.

Estimation of reproduction number

The estimations for daily reproduction number, Re, of SARS-CoV-2 in South Africa were obtained from the covid-19-re data repository (<https://github.com/covid-19-Re/dailyRe-Data>)^2^ as at 15^th^ September 2020. The effective reproductive number describes the average number of secondary infections caused by an infected individual. As described previously^2^, the relevant method of calculation of Re builds upon another method developed by Cori et al.^3^, accessible through EpiEstim R package. Instead of using a time series of infection incidence, which cannot be observed directly, the relevant method infers the infection incidence time series based secondary sources of information such as COVID-19 confirmed case data, hospital admissions, and deaths. This was considered in combination with two other sets of time variables: i) the duration of SARS-CoV-2 incubation period and ii) the time delays between onset of symptoms and a positive test, a hospital admission or the death of a patient. The relevant method infers infection time series from the stated observed incidence data by deconvolution^4,5^.

SARS-CoV-2 samples and metadata

Residual samples from nasopharyngeal and oropharyngeal swabs collected from COVID-19 positive patients obtained from all 11 districts for the province of KwaZulu-Natal (KZN), were used for SARS-CoV-2 WGS. We obtained samples either in the form of primary swabs or extracted RNA. The swab samples were heat inactivated in a water bath at 60°C for 30 minutes, in biosafety level 3 laboratory, prior to RNA extraction. RNA was extracted using the Viral NA/gDNA Kit on the Chemagic 360 system (Perkin Elmer, Hamburg, Germany) using the automated Chemagic 360 insturment (Perkin Elmer, Hamburg, Germany) or manually using the Qiagen Viral RNA Mini Kit (QIAGEN, California, USA). Associated metadata for the samples included date and location (district) of sampling, and sex and age of the patients.

Residual samples from nasopharyngeal and oropharyngeal swabs collected from COVID-19 positive patients obtained from all 11 districts for the province of KwaZulu-Natal (KZN), were used for SARS-CoV-2 WGS. Samples were obtained either in the form of primary swabs or extracted RNA. The swab samples were heat inactivated in a water bath at 60°C for 30 minutes, in biosafety level 3 laboratory, prior to RNA extraction. RNA was extracted using the Viral NA/gDNA Kit using the automated Chemagic 360 system (Perkin Elmer, Hamburg, Germany) or manually using the Qiagen Viral RNA Mini Kit (QIAGEN, California, USA). Associated metadata for the samples included date and location (district) of sampling, and sex and age of the patients.

Real Time RT-PCR

In order to detect the SARS-CoV-2 virus by PCR, the TaqPath COVID-19 CE-IVD RT-PCR Kit (Life Technologies, Carlsbad, CA) was used according to the manufacturer’s instructions. The assays target genomic regions (ORF1ab, S protein and N protein) of the SARS-CoV-2 genome. RT-PCR was performed on a QuantStudio 7 Flex Real-Time PCR instrument (Life Technologies, Carlsbad, CA). Cycle thresholds (Ct) was analysed using auto-analysis settings with the threshold lines falling within the exponential phase of the fluorescence curves and above any background signal.

Whole genome sequencing and genome assembly

cDNA synthesis was performed on the RNA using random primers followed by gene specific multiplex PCR using the ARTIC protocol. Briefly, extracted RNA was converted to cDNA using the Superscript IV First Strand synthesis system (Life Technologies, Carlsbad, CA) and random hexamer primers. SARS-CoV-2 whole genome amplification by multiplex PCR was carried out using primers designed on Primal Scheme (http://primal.zibraproject.org/) to generate 400bp amplicons with an overlap of 70bp that covers the 30Kb SARS-CoV-2 genome. PCR products were cleaned up using AmpureXP purification beads (Beckman Coulter, High Wycombe, UK) and quantified using the Qubit dsDNA High Sensitivity assay on the Qubit 4.0 instrument (Life Technologies Carlsbad, CA).

The Illumina® Nextera Flex DNA Library Prep kit was used according to the manufacturer’s protocol to prepare uniquely indexed paired end libraries of genomic DNA. Sequencing libraries were normalized to 4nM, pooled and denatured with 0.2N sodium acetate. 12pM sample library was spiked with 1% PhiX (PhiX Control v3 adapter-ligated library used as a control). Libraries were loaded onto a 500-cycle v2 MiSeq Reagent Kit and run on the Illumina MiSeq instrument (Illumina, San Diego, CA, USA).

Raw reads coming from Illumina sequencing were assembled using Genome Detective 1.126 (<https://www.genomedetective.com/>) and the Coronavirus Typing Tool ^6,7^. The initial assembly obtained from Genome Detective was polished by aligning mapped reads to the references and filtering out low-quality mutations using bcftools 1.7-2 mpileup method. All mutations were confirmed visually with bam files using Geneious software (Biomatters Ltd, New Zealand). All of the sequences were deposited in GISAID (<https://www.gisaid.org/>)^8^.

Compilation of SARS-CoV-2 South Africa dataset

To present a comprehensive analysis of the genomic epidemiology of SARS-CoV-2 in South Africa, the genomes generated as at 15^th^ September 2020 (n=1111) was combined with all other South African genomes available in GISAID as at the same date (n=298). Appropriate acknowledgement was given to the sequencing laboratories (Supplementary Data S2), and this resulted in a dataset of 1409 genomes. Sampling locations of genomes in this dataset included all provinces in South Africa, and all districts in KZN, the most sampled province, and collection dates spanned from 6^th^ of March 2020 (the first cases in SA) to 26^th^ of August 2020.

Quality control of genome sequences

Prior to phylogenetic reconstruction we filtered out low quality sequences from the dataset. We retrieved all South African SARS-CoV-2 genotypes from the GISAID database as of 26^th^ of August 2020 (N=1,409). We filtered out all genotypes that met any of the following criteria: (1) Sequences with <90% genotype coverage; (2) genotypes with too many mutations (defined as having >20 nucleotide mutations relative to the Wuhan reference); (3) genotypes with >10 ambiguous bases; and (4) genotypes with clustered mutations defined as mutations in close proximity to one another. These are the standard quality assessment parameters employed in NextClade (<https://clades.nextstrain.org>). To this end we analyzed all 1,409 South African genotypes. A total of 16 South African genotypes were filtered out due to low coverage, while a further 28 were removed due to poor sequence quality. The final dataset of South African sequences (N=1365) were further annotated with additional metadata information (sampling locations, unique lab IDs, and outbreak numbers) (Supplementary Figure S9. The bulk of the South African sequences (~81%) were sampled within the province of KZN, with sampling from all of the 11 districts within the province.

Reference global dataset

South African sequences were analyzed against a backdrop of globally representative SARS-CoV-2 genotypes. At the time of sequence analysis, more than 90,000 SARS-CoV-2 genotypes have been publicly shared. Due to the sheer size of this dataset and over sampling and in specific countries (e.g. England) we had to down sample this dataset to a manageable size. Important lineage defining genotypes along with a ten randomly sampled genotypes per location were included in the phylogenetic reconstruction. The final 5,848 references contained 889 other African genotypes, 1,209 genotypes from Asia, 2,775 genotypes from Europe, 434 and 367 genotypes from North and South America respectively and 174 genotypes from Oceania.

Phylogenetic analysis of SARS-CoV-2 in South Africa

South African genotypes were analyzed against the global reference dataset using a custom build of the SARS-CoV-2 NextStrain build (https://github.com/nextstrain/ncov). The pipeline contains several python scripts that manage the analysis workflow. In short it allows for the filtering of genotypes, the alignment of genotypes in MAFFT^9^, phylogenetic tree inference in IQ-Tree^10^, tree dating and ancestral state construction and annotation. The resulting time scaled phylogeny can be viewed interactively and has been shared publicly on the NGS-SA NextStrain page (<https://nextstrain.org/groups/ngs-sa/COVID19-Africa-2020.09.16>).

The raw ML-tree topology that was produced by the NextStrain build were used to estimate the number of viral introductions through time into South Africa. TreeTime^11^ was used to transform this ML-tree topology into a dated tree topology using a constant rate of 8.0 x 10^-4^ nucleotide substitutions per site per year, following the exclusion of outlier sequences. A migration model was fitted on the resulting time scaled tree topology in TreeTime mapping country locations to tips and internal nodes. The resulting annotated tree topology was used to infer the number of viral introductions into South Africa through time.

Lineage & Clade classification

We used the dynamic lineage classification method proposed by Rambault et al.^12^ in this study via the Phylogenetic Assignment of named Global Outbreak LINeages (PANGOLIN) software suite (<https://github.com/hCoV-2019/pangolin>). This is aimed at identifying the most epidemiologically important lineages of SARS-CoV-2 at the time of analysis, allowing researchers to monitor the epidemic in a particular geographical region. Three main SARS-CoV-2 lineages are currently recognized; lineage A, defined by Wuhan/WH04/2020, lineage B, defined by Wuhan-Hu-1 strain, and lineage C, a sub-classification from the B lineage. We also classified the SARS-CoV-2 genomes in our dataset using the clade classification proposed by Nextstrain, divided into 19A, 19B, 20A, 20B, and 20C clades^13,14^.

Dated phylogenetics

To estimate time-calibrated phylogenies dated from time-stamped genome data, we conducted phylogenetic analysis using the Bayesian software package BEASTv.1.10.4^15^, on three smaller subsets of data for each of the three lineages identified in the ML phylogeny and containing isolates from South Africa (Cluster B.1.1.54, n = 320; Cluster B.1.1.56, n = 104; Cluster C.1, n = 151; Cluster B.1.106, n = 68).

ML trees from these three data subsets were inspected in TempEst v1.5.3 for the presence of a temporal (i.e. molecular clock) signal^16^. Linear regression of root-to-tip genetic distances against sampling dates indicated that the SARS-CoV-2 sequences evolve in a relatively-strong clock-like manner (r = 9.45e-2; r = 0·34; r=0.74, r = 0·50 from subset B.1.1.54; B.1.1.56 and C.1, respectively) (Supplementary Figure S5).

For this analysis we employed the strict molecular clock model, the HKY+I, nucleotide substitution model and the exponential growth coalescent model^17^. We computed MCMC (Markov chain Monte Carlo) triplicate runs of 100 million states each, sampling every 10.000 steps for each data set. Convergence of MCMC chains was checked using Tracer v.1.7.1^18^. Maximum clade credibility trees were summarised from the MCMC samples using TreeAnnotator after discarding 10% as burn-in.

Phylogeographic analysis

To model phylogenetic diffusion of South African lineages across the country, we used a flexible relaxed random walk (RRW) diffusion model that accommodates branch-specific variation in rates of dispersal with a Cauchy distribution^19^. For each sequence, latitude and longitude were attributed to a point randomly sampled within the patient’s province or district of residence. We discretised sequence sampling locations by considering 5 of 9 provinces in South Africa, and all 11 districts in KZN, the most sampled province, where sequences belonging to the three cluster were sampled (as shown Supplementary Fig S3).

MCMC chains were run for >100 million generations and sampled every 10000th step, with convergence assessed using Tracer v1.7^20^. Maximum clade credibility trees were summarized using TreeAnnotator after discarding 10% as burn-in. We used the R package “seraphim”^21,22^ to extract and map spatiotemporal information embedded in posterior trees.

6.	Vilsker, M. *et al.* Genome Detective: an automated system for virus identification from high-throughput sequencing data. doi:10.1093/bioinformatics/bty695.

7.	Cleemput, S. *et al.* Genome Detective Coronavirus Typing Tool for rapid identification and characterization of novel coronavirus genomes. doi:10.1093/bioinformatics/btaa145.

8.	Jasper J. Koehorst, Jesse C. J. van Dam, Edoardo Saccenti, Vitor A. P. Martins dos Santos, M. S.-D. and P. J. S. GISAID Global Initiative on Sharing All Influenza Data. Phylogeny of SARS-like betacoronaviruses including novel coronavirus (nCoV). *Oxford* **34**, 1401–1403 (2017).

9.	Katoh, K. & Standley, D. M. MAFFT multiple sequence alignment software version 7: improvements in performance and usability. *Mol. Biol. Evol.* **30**, 772–780 (2013).

10.	Nguyen, L.-T., Schmidt, H. A., Von Haeseler, A. & Minh, B. Q. IQ-TREE: A Fast and Effective Stochastic Algorithm for Estimating Maximum-Likelihood Phylogenies. doi:10.1093/molbev/msu300.
