## Supplementary Figures for "Major new lineages of SARS-CoV-2 emerge and spread in South Africa during lockdown"

**Supplementary Section 2 – Supplementary Figures**

**Contents:**

- Supplementary Figures S1 to S9
- Supplementary Tables S1
- Captions for Data S1, Data S2, and Data S3

**Additional Supplementary materials:**

- Materials and Methods
- Data S1 – South Africa dataset metadata (Excel file)
- Data S2 – Acknowledgements to sequence-generating laboratories in South Africa dataset (PDF file)
- Data S3 – Acknowledgements to sequence-generating laboratories in global reference dataset (Excel file)

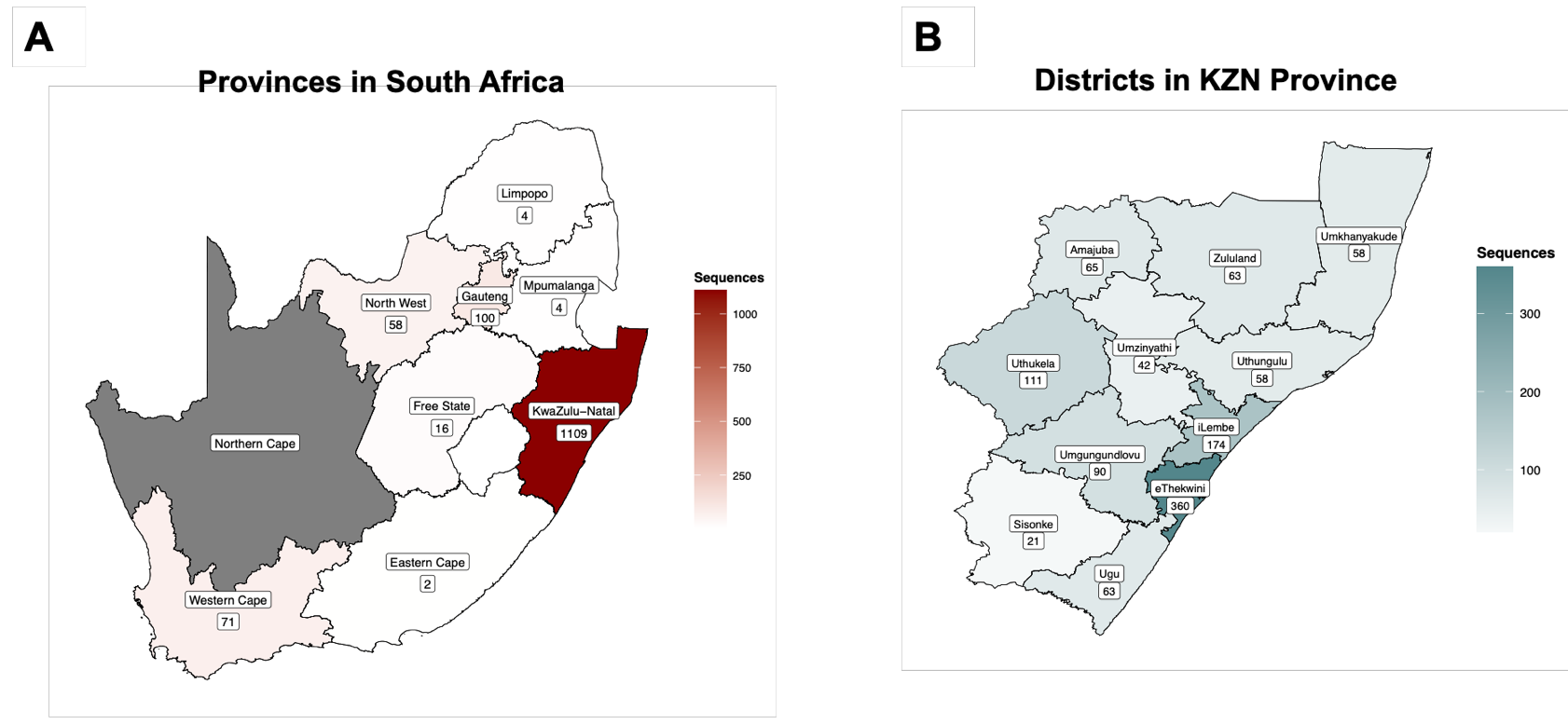

**Supplementary Figure S1** – Map density representation of where the genomes in this study were sampled, showing the number of genomes sampled in each province in South Africa (left) (no genomes from Northern Cape – grey) and the number of genomes sampled in each district of KZN, the most sampled province (right).

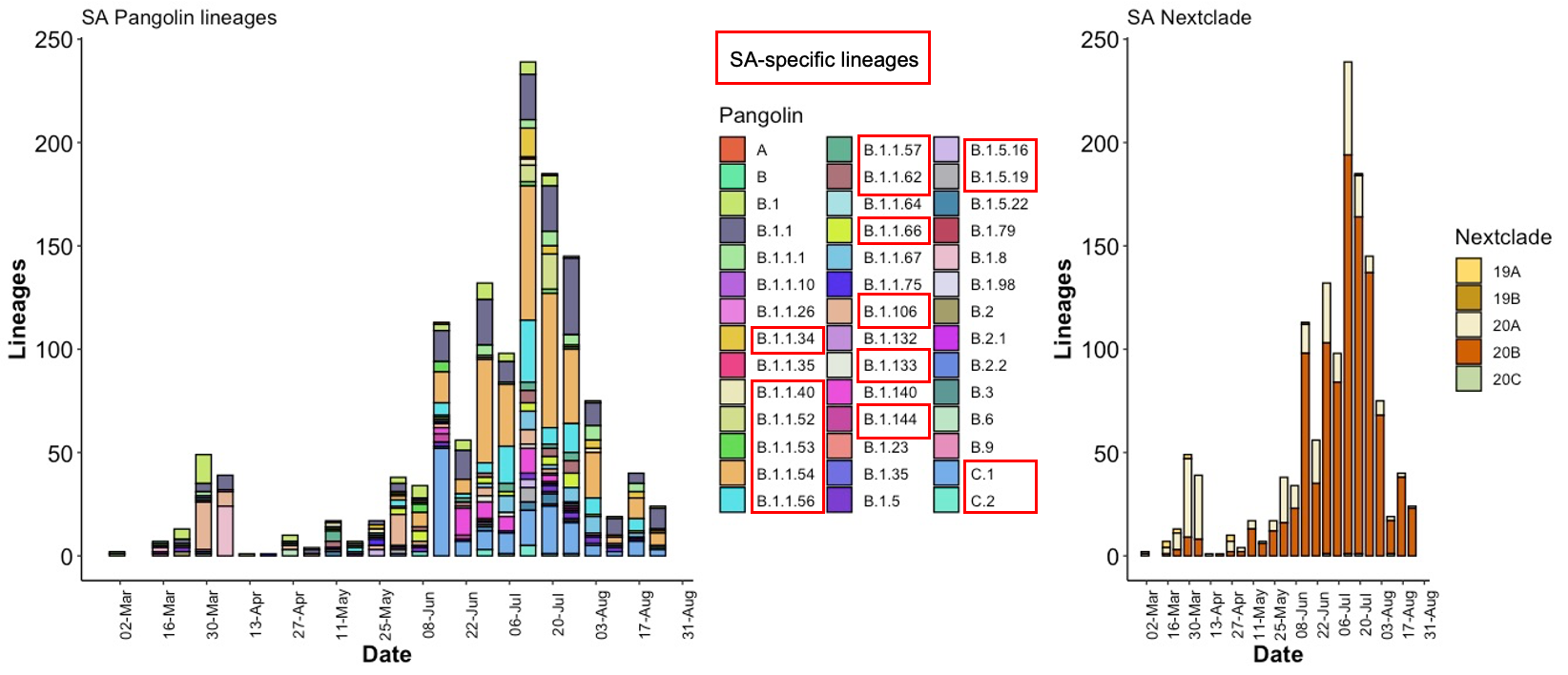

**Supplementary Figure S2** – Classification of South Africa genomes (n=1365) per date into Pangolin lineages (SA-specific ones specified by red boxes), and into Nextstrain clades.

**Supplementary Table 1:** Detailed sampling information for the four lineage cluster identified to be almost unique to South Africa

|  | B.1.106 | B.1.1.54 | C.1 | B.1.1.56 |
| --- | --- | --- | --- | --- |
| Number of Genomes in our dataset | 68 | 320 | 151 | 104 |
| Most common countries | ZA (56%), USA (39%), Spain (3%) | ZA (99%), UK (1%) | ZA (99%), UK (1%) | ZA (99%), Australia (1%) |
| Most Common Provinces | KwaZulu-Natal | North West, Gauteng, KwaZulu-Natal | North West, Gauteng, Limpopo, Free State, KwaZulu-Natal | KwaZulu-Natal |
| KZN districts | 5 districts | All 11 districts | All 11 districts | All 11 districts |
| Date range | March 16 to July 21 | March 19 to August 26 | June 03 to August 26 | March 21 to August 21 |
| Dates since last sampling in ZA | 2020-08-21 | 2020-08-26 | 2020-08-21 | 2020-08-26 |
| Recall value | 0.84 | 0.98 | 0.99 | 0.95 |

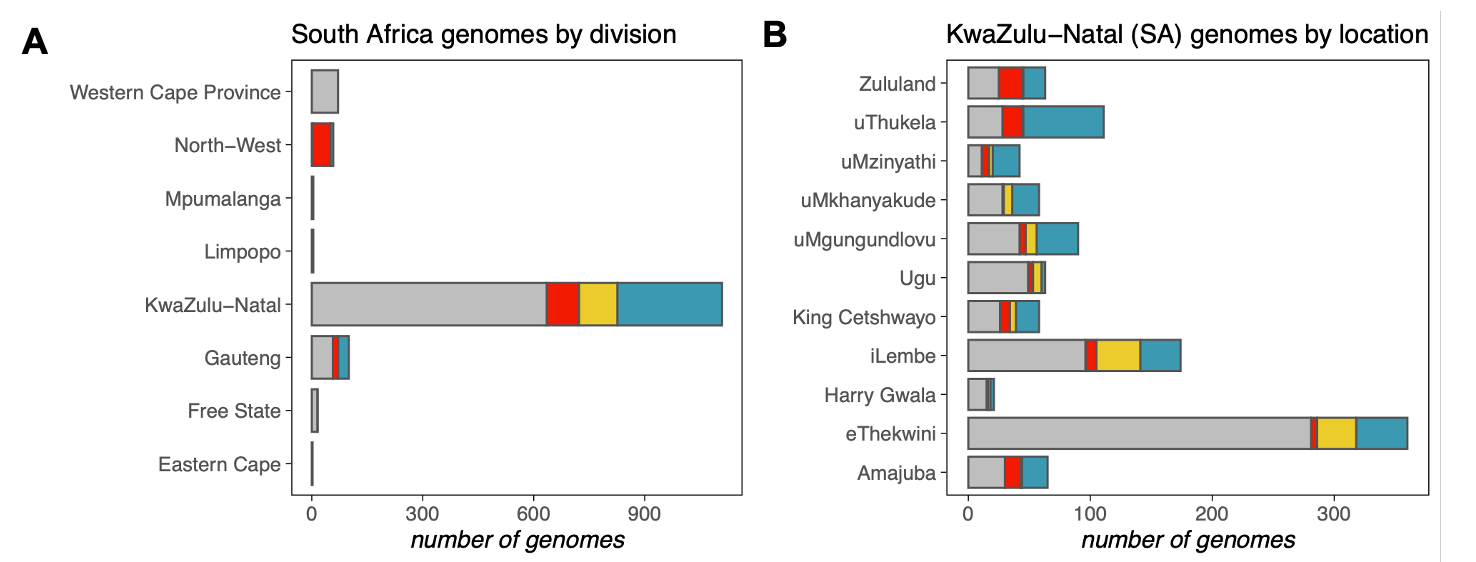

**Supplementary Figure S3**. A) Distribution of genomes belonging to the lineage clusters by province. B) Distribution of genomes belonging to the lineage clusters by district of KZN.

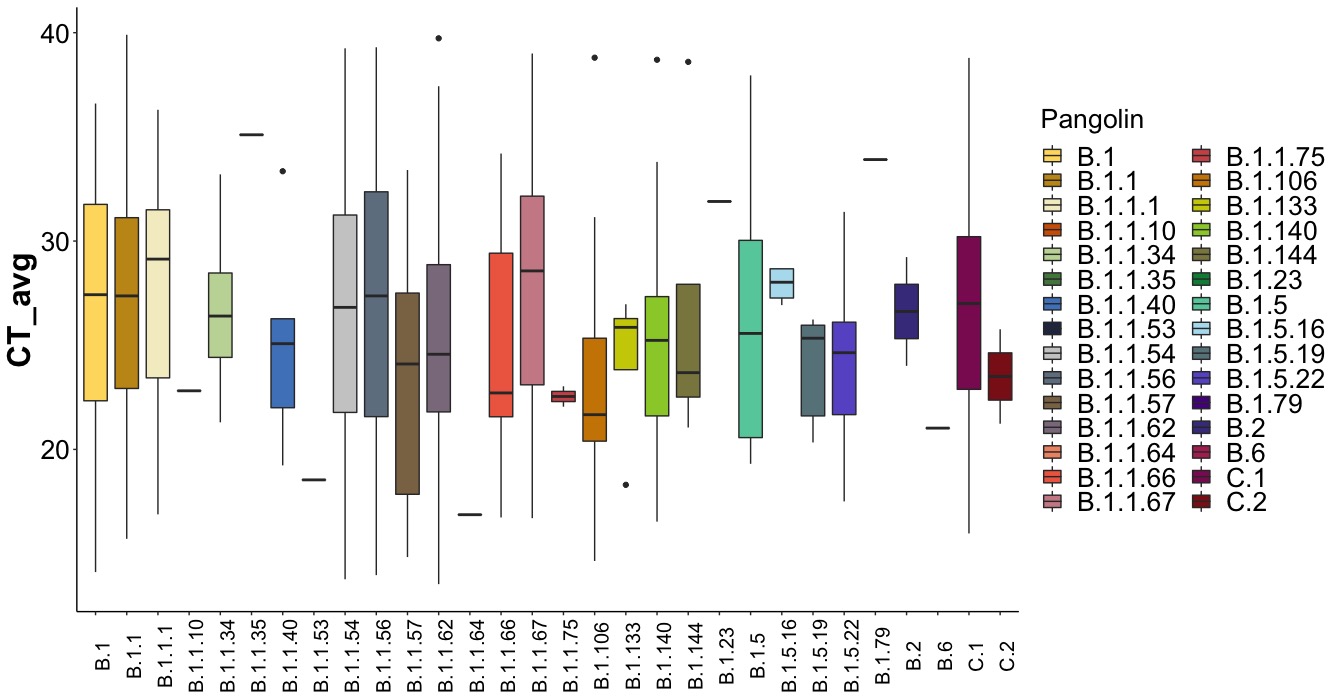

**Supplementary Figure S4** – Showing the average Ct scores at three target genes for genomes generated at KRISP, and classified into their respective Pangolin lineages

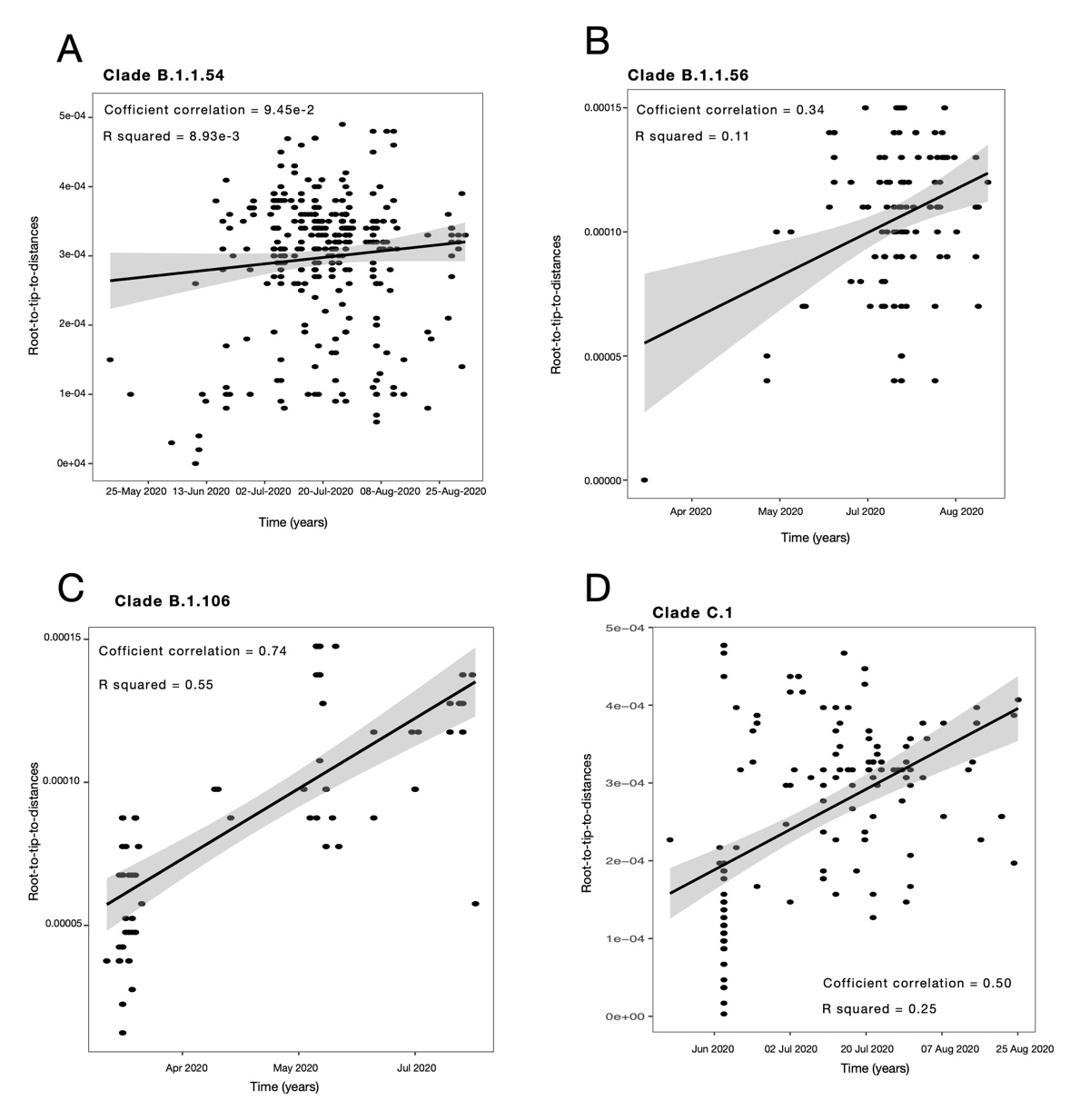

**Supplementary Figure S5-** Temporal signaling for each cluster (Tempest). For SARS-CoV-2, we accept temporal signaling with correlation coefficient > 0.2. Cluster B.1.1.54 (A) had a low correlation coefficient and was therefore rejected from further Bayesian spatiotemporal analyses.

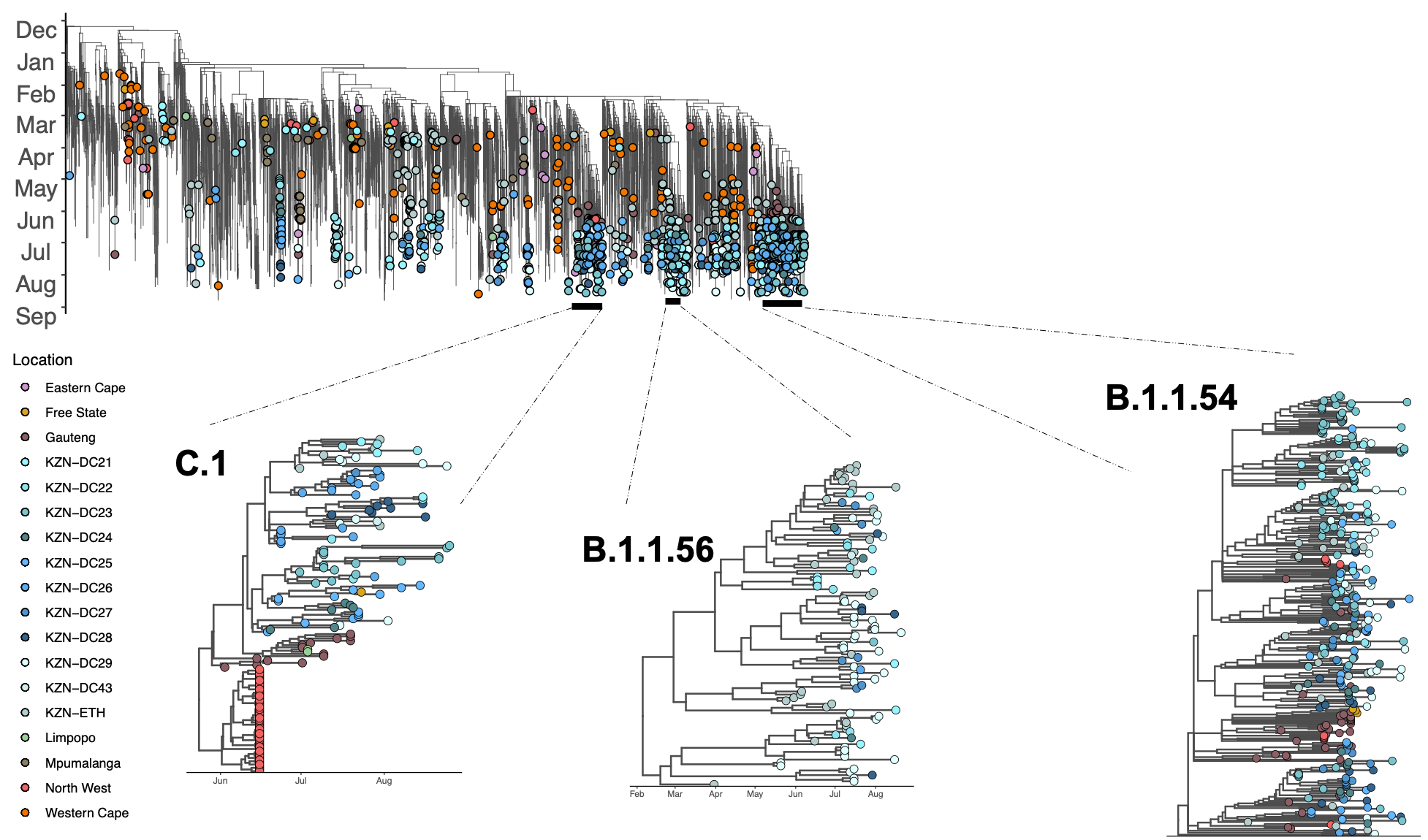

**Supplementary Figure S6.** Maximum likelihood tree of a global dataset showing genomes coloured by sampling location in South Africa. For genomes sampled in KZN, they are further specified by which district they were sampled from. A closer look into cluster B.1.106, C.1 and B.1.1.56 illustrated as trees from BEAST temporal analyses, with a defined time-scale. The zoom-in tree for B.1.1.54 was extracted as a subset of the big ML tree.

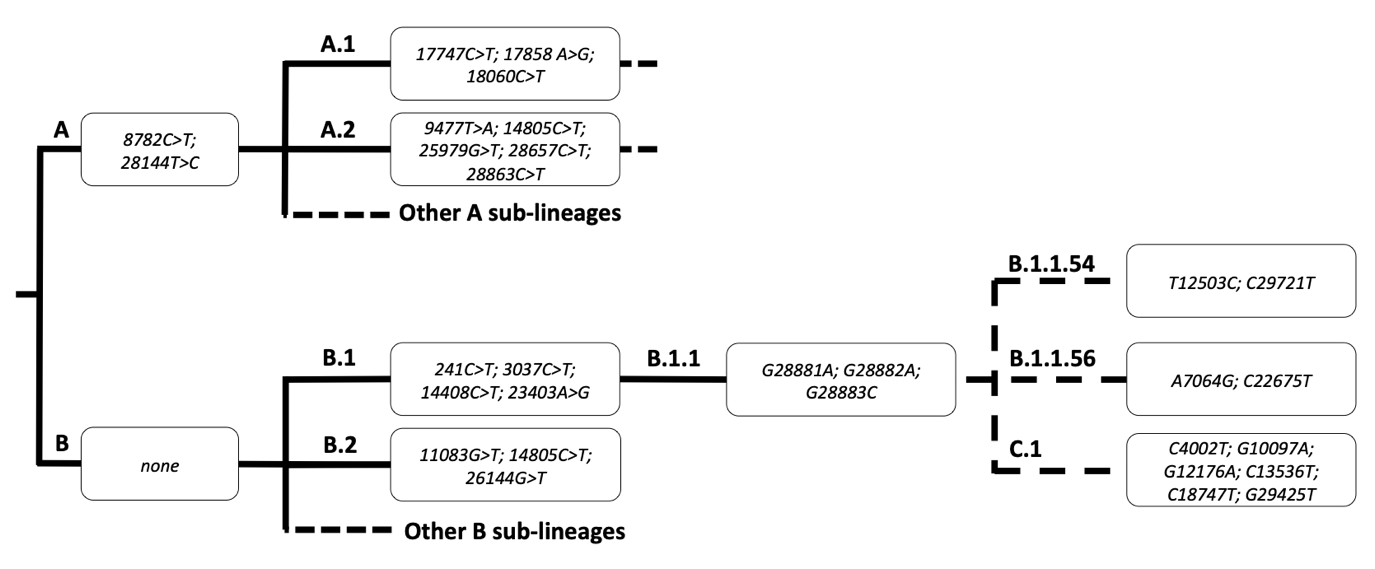

**Supplementary Fig S7**. Flowchart of Pangolin lineage A and B dividing into sub-lineages with their lineage-defining mutations specified

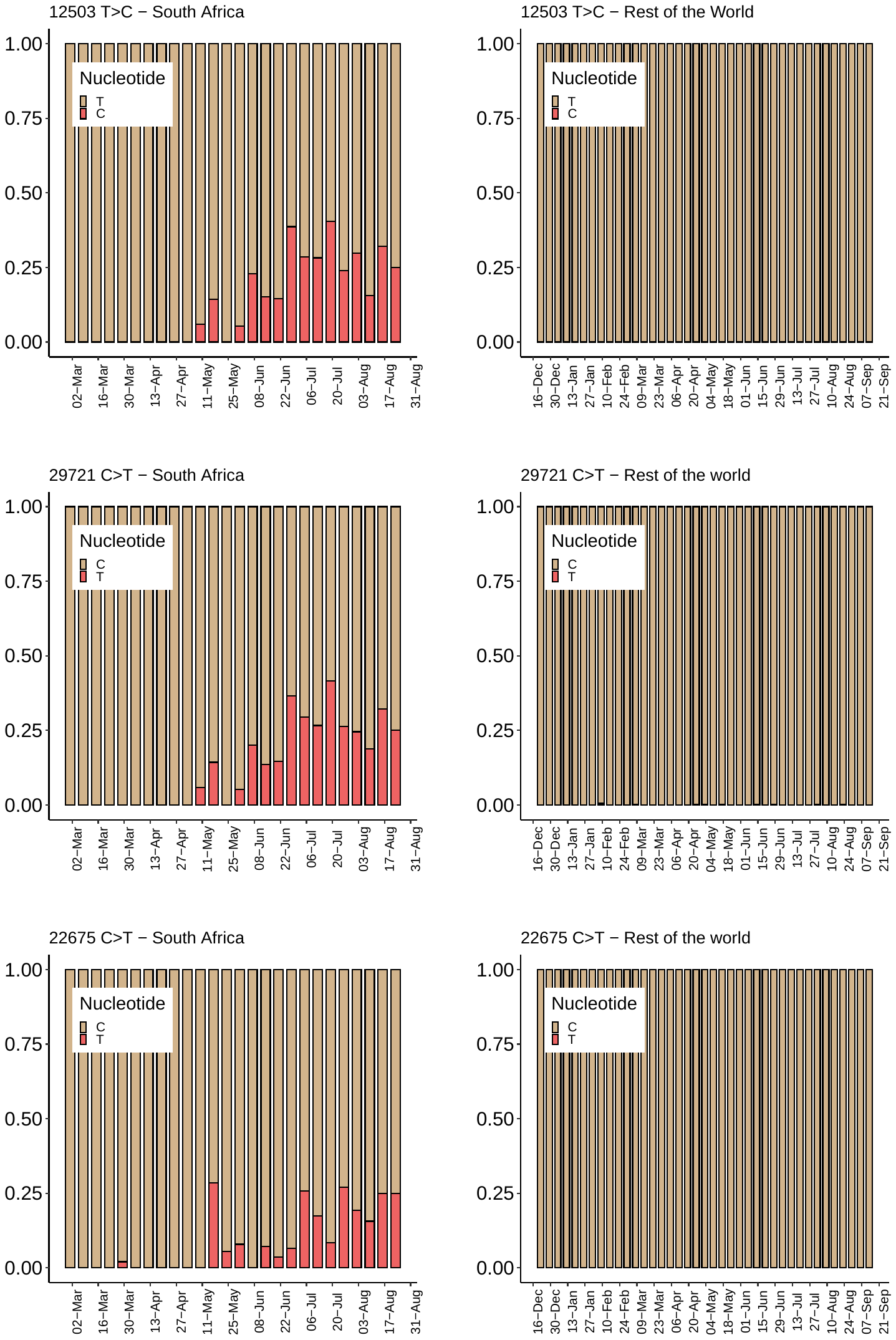

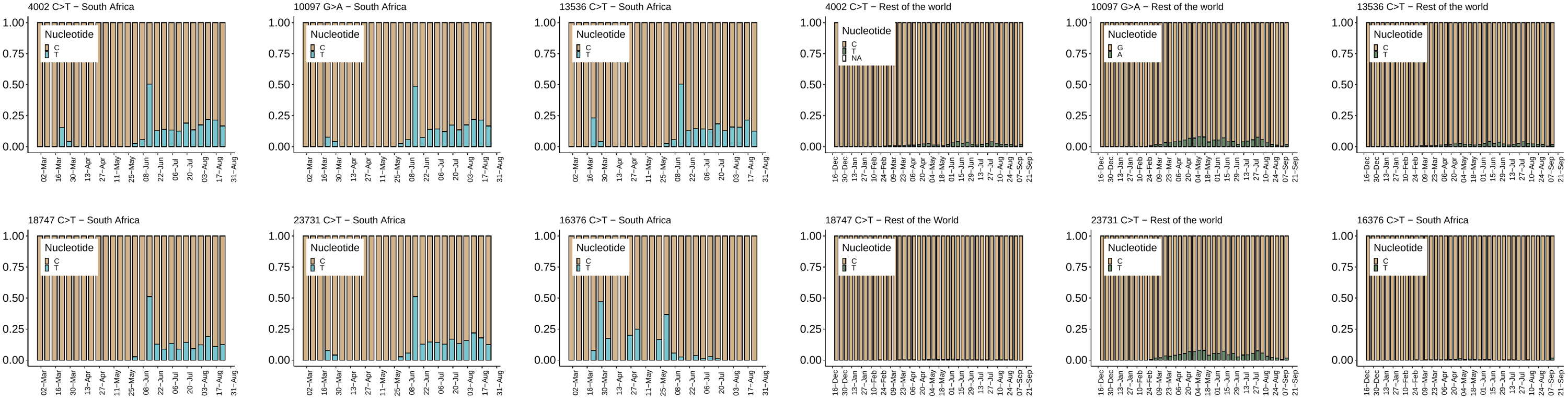

**Supplementary Fig S8** – Mutation frequencies in SA vs rest of the world for lineage-defining mutations

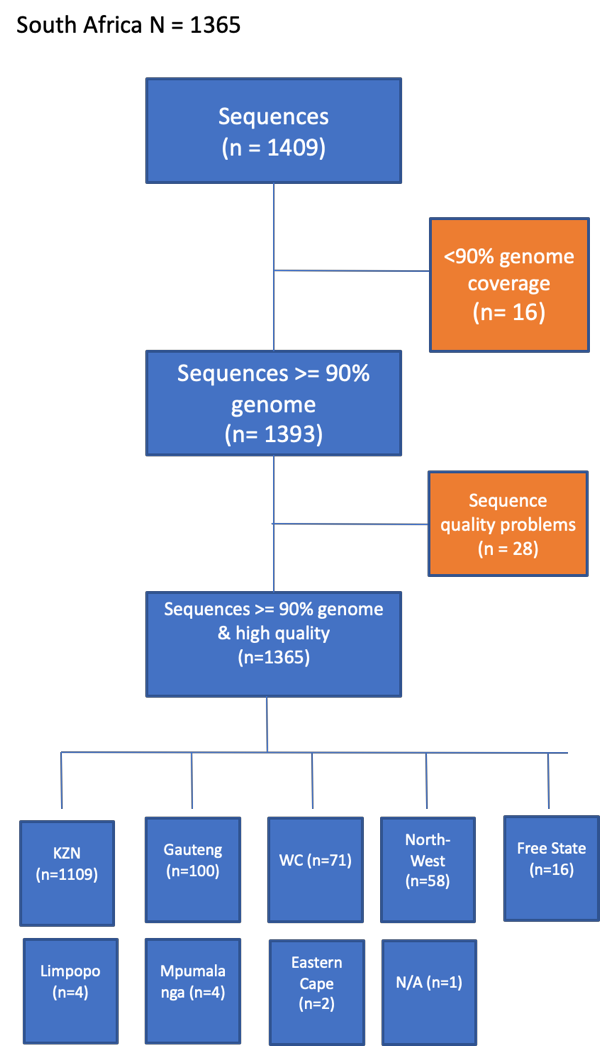

**Supplementary Fig S9 –** Curation of South Africa dataset from all available South African genomes available on GISAID as at 15^th^ September 2020, showing the initial number of genomes (n=1409), how many were excluded at each cleaning step and the final number of genomes (n=1365) with subdivisions into their origininating province.

**Captions for Data S1, Data S2, and Data S3**

Data S1: Metadata table for 1365 South African SARS-CoV-2 sequences available on GISAID as at 15^th^ September 2020.

Data S2: Acknowledgement to all laboratories in South-Africa responsible for producing SARS-CoV-2 genomes that were the focus of the analysis in this paper

Data S3: Acknowledgement table for all global SARS-CoV-2 sequences (obtained through GISAID) used as reference dataset in this study.
